# Exponential phase of covid19 expansion is driven by airport connections

**DOI:** 10.1101/2020.04.02.20050773

**Authors:** Marco Túlio Pacheco Coelho, João Fabricio Mota Rodrigues, Anderson Matos Medina, Paulo Scalco, Levi Carina Terribile, Bruno Vilela, José Alexandre Felizola Diniz-Filho, Ricardo Dobrovolski

## Abstract

The pandemic state of COVID-19 caused by the SARS CoV-2 put the world in quarantine, led to hundreds of thousands of deaths and is causing an unprecedented economic crisis. However, COVID-19 is spreading in different rates at different countries. Here, we tested the effect of three classes of predictors, i.e., socioeconomic, climatic and transport, on the rate of daily increase of COVID-19. We found that global connections, represented by countries’ importance in the global air transportation network, is the main explanation for the growth rate of COVID-19 in different countries. Climate, geographic distance and socioeconomics had a milder effect in this big picture analysis. Geographic distance and climate were significant barriers in the past but were surpassed by the human engine that allowed us to colonize most of our planet land surface. Our results indicate that the current claims that the growth rate of COVID-19 may be lower in warmer and humid tropical countries should be taken very carefully, at risk to disturb well-established and effective policy of social isolation that may help to avoid higher mortality rates due to the collapse of national health systems.

## Introduction

With the worldwide spread of the novel Coronavirus Disease 2019 (COVID-19), caused by the SARS-CoV-2 virus, we are experiencing a declared pandemic. One of the largest preoccupations about this new virus regards its notable ability to spread given the absence of any effective treatment, vaccine and immunity in human populations. Epidemiologists quantify the ability of infectious agents to spread by estimating the basic reproduction number (*R0*) statistic [1], which measures the average number of people each contagious person infects. According to the World Health Organization [2], the new coronavirus is transmitting at an *R0* around *1.4-2.5,* which is greater than seasonal influenza viruses that spread every year around the planet (median *R0* of 1.28, [3]). To anticipate the timing and magnitude of public interventions and mitigate the adverse consequences on public health and economy, understanding the factors associated with the survival and transmission of SARS-CoV-2 is urgent.

Because previous experimental [4], epidemiological [5, 6] and modeling [7] studies show the critical role of temperature and humidity on the survival and transmission of viruses, recent studies are testing the effect of environmental variables on SARS-CoV-2 [8, 9] and forecasting monthly scenarios of the spread of the new virus based on climate suitability [10, *but see* 11]. Although temperature and humidity are known to affect the spread and survival of other coronaviruses (i.e., SARS-CoV and MERS-CoV, [12–15] using the current occurrences of SARS-CoV-2 cases to build correlative climatic suitability models without taking into consideration connectivity among different locations and socioeconomic conditions might be inadequate.

Many factors might influence the distribution of diseases at different spatial scales. Climate might affect the spread of viruses given its known effect on biological processes that influences many biogeographical patterns, including the distribution of diseases and human behavior (e.g., [16]). Geographic distance represents the geographical space where the disease spread following the distribution of hosts and has also been found to explain biogeographic patterns [17–19]. Socioeconomic characteristics of countries could be viewed as a proxy for the ability to identify and treat infected people and for the governability necessary to make fast political decision and avoid the spread of new diseases. Finally, the global transportation network might surpass other factors as it can reduce the relative importance of geographic distance and facilitate the spread of viruses and their vectors [20,21]. According to the International Air Transport Association [22] more than 4 billion passengers traveled abroad in 2018. This amount of travelers reaching most of our planet’s surface represents a human niche construction (i.e. global transportation network; [23]) that facilitates the global spread of viruses and vectors [20,21] in the same way it facilitated the spread of invasive species and domesticated animals over modern human history [23].

The spread of SARS-CoV-2 from central China to other locations might be strongly associated with inter-country connections, which might largely surpass the effect of climate suitability. Thus, at this point of the pandemic, there is still a distributional disequilibrium that can generate very biased predictions based on climatic correlative modeling [24]. Thus, here we used an alternative macroecological approach [25], based on the geographical patterns of exponential growth rates of the disease at country level, to investigate variations on the growth rates of SARS-CoV-2. We studied the effect of environment, socioeconomic and global transportation controlling for spatial autocorrelation that could bias model significance. By analyzing these factors, we show that the exponential growth of COVID-19 at global scale is explained mainly by country’s importance in global transportation network (i.e., air transportation).

## Material and methods

We collected the number of people infected by the COVID-19 per day from the John Hopkins [26] and European Centre for Disease Prevention and Control (ECDC, [27]). This data is available for 204 countries, for which 65 had more than 100 cases recorded and for which time series had at least 30 days after the 100^th^ case. We also performed the analysis considering countries with more than 50 cases, but it did not qualitatively change our results. Thus, we only show the results for countries with more than 100 cases.

In our analysis, we only used the exponential portion of the time series data (i.e. number of people infected per day) and excluded days after stabilization or decrease in total number of cases. We empirically modelled each time series using an exponential growth model for each country and calculated both the intrinsic growth rate (r) and the regression coefficient of the log growth series to be used as the response variable in our models. Because both were highly correlated (Person’s r = 0.97), we used only the regression coefficient to represent the growth rate of COVID-19 in our study.

To investigate potential correlates of the virus growth rate, we downloaded climatic and socioeconomic data of each country. We used climatic data represented by monthly average minimum and maximum temperature (°C) and total precipitation (mm) retrieved from the WorldClim database (https://www.worldclim.org) [29]. We used monthly available data for the most recent year available in WorldClim. We extracted climatic data from the months of January, February, March, and December to represent the climatic conditions of the winter season in the Northern Hemisphere and the summer season in the Southern Hemisphere. From these data, we computed the mean value of climatic variables across each country. Finally, minimum and maximum temperatures were combined to estimate monthly mean temperature for December, January, February, and March, which was used in the model along with total precipitation for the same months. However, using different combinations of these variables (i.e., using means of minimum or maximum temperatures, as well as minimum or maximum for each month) did not qualitatively affect our results.

We extracted socioeconomic data for each country. Human Development Index (HDI) rank, mean number of school years in 2015, gross national income (GNI) per capita in 2011, population size in 2015 and average annual population growth rate between 2010-2015 were used in our study and downloaded from the United Nations database (http://hdr.undp.org/en/data). We also obtained a mean value of health investment in each country by averaging the annual health investments between 2005 2015 obtained from the World Health Organization database (http://apps.who.int/gho/data/node.home). Due to the strong collinearity among some of these predictors, HDI rank and mean number of school years were removed from our final model.

Finally, we also downloaded air transportation data from the OpenFlights [30] database regarding the airports of the world, which informs where each airport is located including country location (7,834 airports), and whether there is a direct flight connecting the airports (67,663 connections). We checked the Openfligths database to make the airports and connections compatible by including missing or fixing airport codes and removing six unidentified airport connections resulting in a total of 7,834 airports and 67,657 connections. We used this information to build an air transportation network that reflects the existence of a direct flight between the airports while considering the direction of the flight. Thus, the airport network is a unipartite, binary, and directed graph where airports are nodes and flights are links (Fig 1, Fig S1). In the following step, we collapsed the airports’ network into a country-level network using the country information to merge all the airports located in a country in a single node (e.g., United States had 613 airports that were merged in a single vertex representing the country). The country-level network (Fig 1, Fig S1) is a directed weighted graph where the links are the number of connections between 226 countries which is collapsed for the 65 countries that had more than 100 cases and for which time series data had at least 30 days after the 100^th^ case. Afterward, we measured the countries centrality in the network using the Eigenvector Centrality [31], hereafter centrality, that weights the importance of a country in the network considering the number of connections with other countries and how well connected these countries are to other countries – indirect connections. All networks analyses were generated using package *igraph* [32].

**Fig 1.**
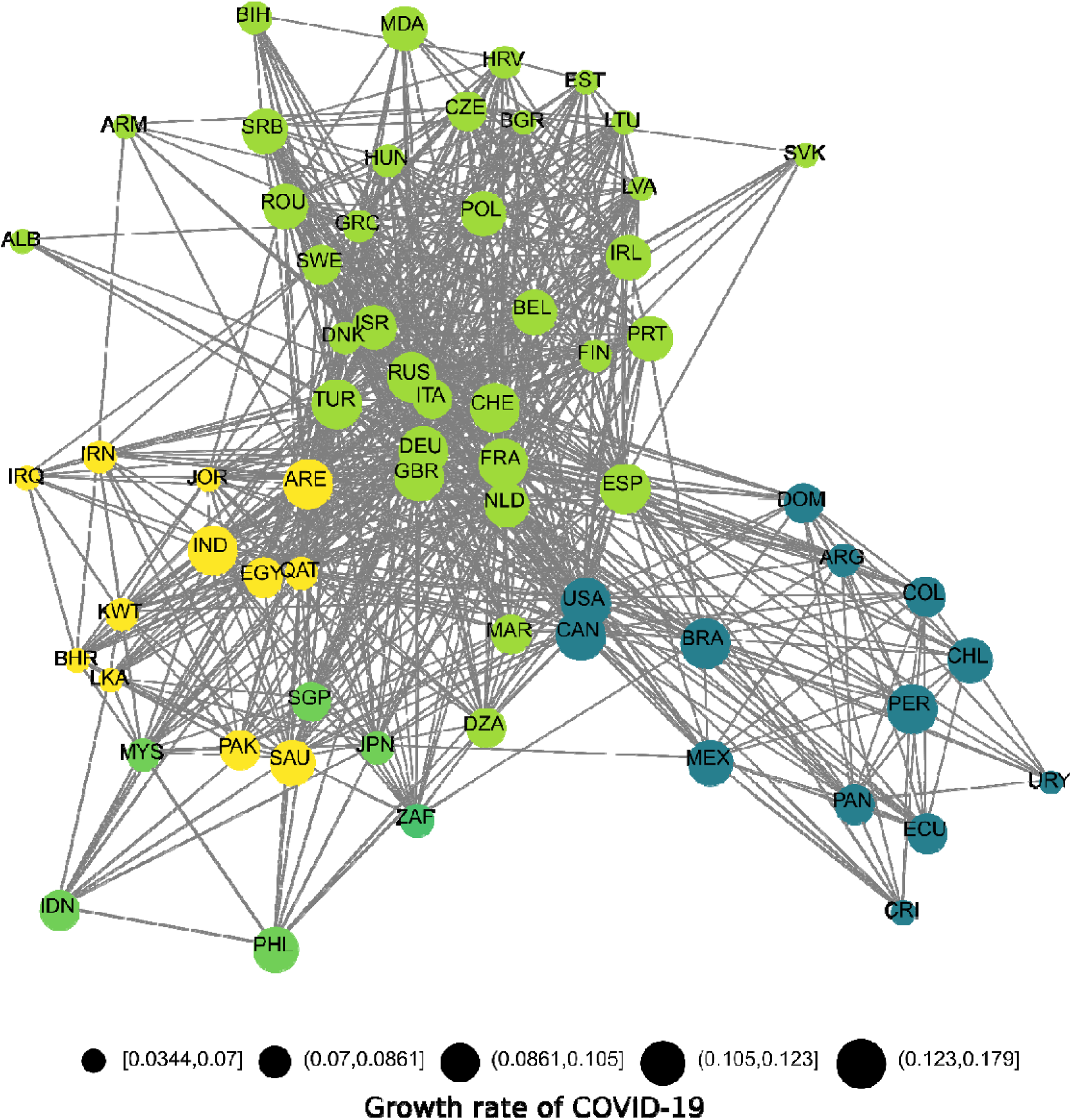
Air transportation network among 65 countries that had more than 100 cases and for which time series data had at least 30 days after the 100^th^ case. Different colours represent modules of countries that are more connected to each other. Different sizes of each node represent the growth rate of COVID-19 estimated for each country (See results Fig 2).

We evaluated the relationship between the predictors (climatic, socioeconomic and transport data) and our growth rate parameter using a standard multiple regression (OLS) after taking into consideration the distribution of the original predictors as well as the normality of model residuals. Moreover, OLS residuals were inspected to evaluate the existence of spatial autocorrelation that could upward bias the significance of predictor variables on the model using Moran’s I correlograms [33]. Prior to the analysis, we applied logarithmic (mean precipitation, total population size, and network centrality) and square root (mean health investments) transformations to the data to approximate a normal distribution.

## Results

The models used to estimate COVID-19 growth rate on different countries showed an average R^2^ of 0.91 (SD = 0.04), varying from 0.78 to 0.99, indicating an overall excellent performance on estimating growth rates. The geographical patterns in the growth rates of COVID-19 cases do not show a clear trend, at least in terms of latitudinal variation, that would suggest a climatic effect at macroecological scale (Fig. 2A).

**Fig 2.**
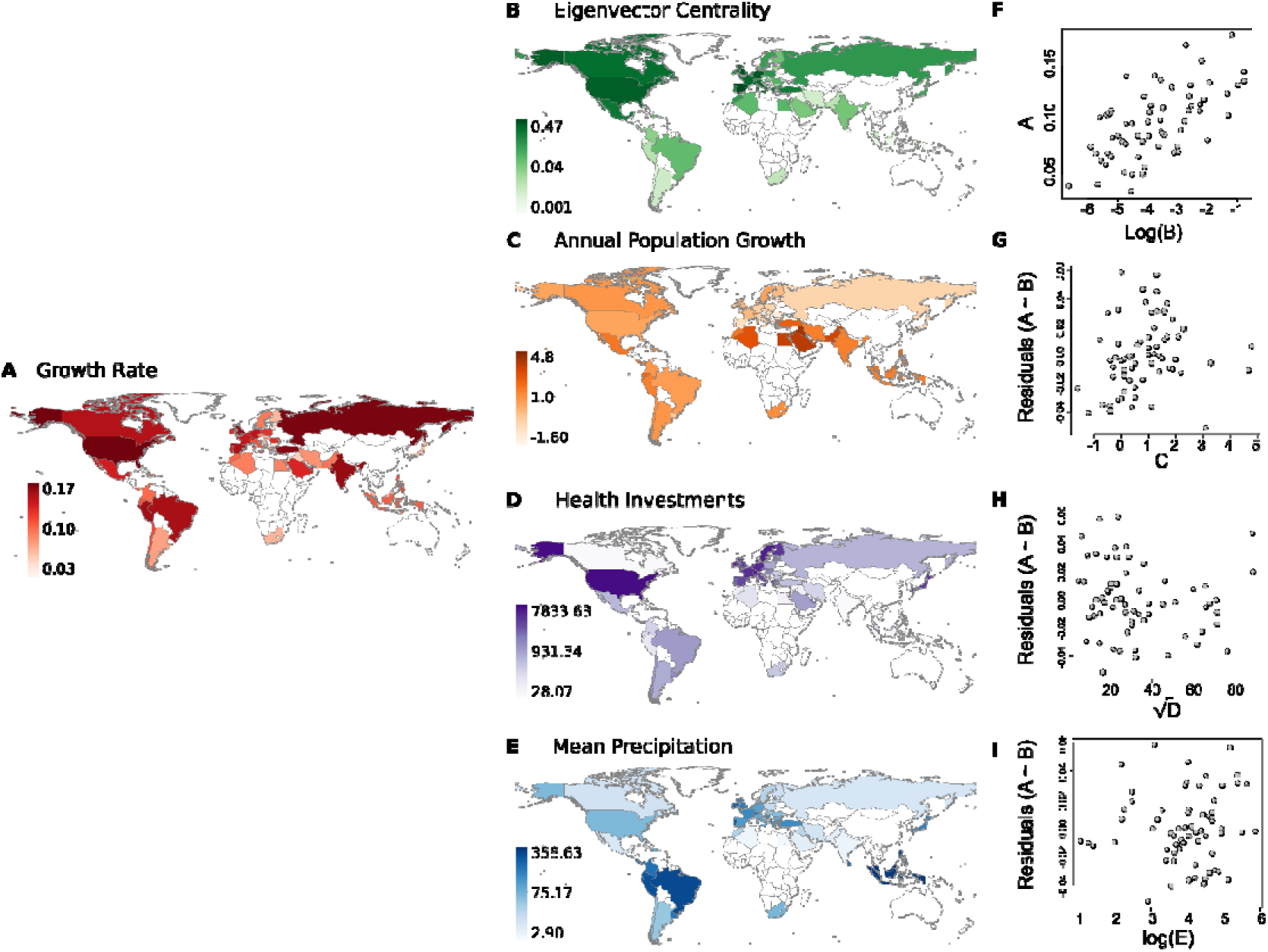
Geographical patterns of growth rate of covid-19 in the exponential phase **(A)**, the Eigenvector Centrality that represents countries’ importance in global transportation network **(B)**, Annual population growth **(C)**, health investments **(D)** and mean precipitation **(E)**. The relationship between growth rate and the log transformed eigenvector centrality is showed in **F. G, H** and **I** are partial plots showing the relationship between the residuals of growth rates vs log transformed eigenvector centrality and annual population growth, health investments and mean precipitation.

We build one model including only climate and socioeconomic variables, which explained *only 14%* of the variation on growth rates. This model did not have spatial autocorrelation in the residuals. When we added country centrality (i.e. country importance in global transportation network) as a predictor, the R^2^ increased to 48.6%. In this model, annual population growth and precipitation had positive and significant effects (Table 1, P = 0.036, P =0.041, Fig 2), while health investments had a negative and significant effect on growth rate (Table 1, P= 0.035, Fig 2). Here, exponential growth rates increased *strongly* in response to countries importance in the transportation network which has more than two times the effect size of any significant variable (Table 1). However, it is also important to note that growth rates of COVID-19 weakly increases with increases of annual population growth and precipitation, and decreases with higher investments in health (Table 1). Statistical coefficients were not upward biased by spatial autocorrelation.

**Table 1.** Model statistics for all variables used in the study.

|  | Standardized |  |  |  |  |
| --- | --- | --- | --- | --- | --- |
|  | Estimate | Estimate | Std Error | t value | P-value |
| *Intercept* |  | 0.149 | 0.026 | 3.547 | < 0.001 |
| *Eigenvector Centrality* | 0.758 | 0.016 | 0.002 | 6.124 | < **0.001** |
| *Gross National Income* | 0.16 | 0.000 | 0.000 | 1.491 | 0.141 |
| *Population Size* | -0.096 | -0.004 | 0.004 | -0.953 | 0.344 |
| *Annual population growth* | 0.306 | 0.008 | 0.003 | 2.139 | **0.036** |
| *Heath investment* | -0.287 | -0.0004 | 0.000 | -2.155 | **0.035** |
| *Mean Temperature* | -0.127 | -0.0003 | 0.000 | -0.908 | 0.367 |
| *Mean Precipitation* | 0.253 | 0.007 | 0.003 | 2.091 | **0.041** |

## Discussion

The pandemic state of SARS CoV-2 is killing hundreds of thousands of people, put the world in quarantine and is causing an unprecedented economic crisis. The rates of increase of new cases of COVID-19 is faster in some countries than others. To understand why growth rates are different among countries we investigate the effect of climatic, socioeconomical and human transportation variables that could have important roles on the exponential phase of COVID-19. At global scale, temperature, population size and Gross National Income had no significant effect on the exponential phase of COVID-19. However, annual population growth, health investments and precipitation show significant, but weak effects on growth rates. Countries’ importance in the global transportation network has a key role on the severity of COVID-19 pandemic in different countries as it is strongly associated with the growth rates of the disease (Fig 2).

The centrality measure is widely used to discover distinguished nodes on many networks, including epidemiological networks (e.g., [34]). Our findings reinforce the importance of propagule pressure on disease dissemination [35, 36]. Aerial transportation is an important predictor of COVID-19 dissemination in China [37], in Brazil [38], and in Mexico [39]. It is quite likely that further phases of COVID-19 spread, in terms of peak of infections and decrease in mortality rates, will be better related to socioeconomics characteristics of each county and their political decisions when secondary transmissions were identified. We can already clearly identify the effects of adopting strong social isolation policies in China (see [37]) and, on the opposite side of this spectrum, in European countries like Italy, Spain and England [40]. Our analyses call attention to the case of Brazil, a well-connected tropical country that presents one of the highest increase rates of COVID-19 in the tropics in its exponential phase (Fig 2A). If decision makers take into consideration yet unsupported claims that growth rates of COVID-19 in its exponential phase might be lower in warmer and humid countries, we might observe terrible scenarios unrolling in tropical countries, especially in those with limited health care structure, such as Brazil. As our results also show, those countries that invested less in health are also the ones with faster growth rates of covid-19 in its exponential phase (Fig 2, Table 1).

When discussing and modelling the effect of climate on SARS CoV-2 it is important to remember that modern human society is a complex system composed of strongly connected societies that are all susceptible to rare events. It is also critical to consider the negative correlations between climate and local or regional socioeconomic conditions (i.e., inadequate sanitary conditions and poor nutritional conditions) that could easily counteract any potential climatic effect at local scales, such as lower survival rates of viruses exposed to high humidity, temperatures and high UV irradiation [8, 41] Tropical regions will experience mild climate conditions in a couple of months. Thus, regardless of the influence of local environmental conditions, tropical countries could still expect high contagious rates. In addition, our results points to a positive effect of precipitation on growth rates, which is the contrary of what has been suggested by climate suitability models. Finally, climatic suitability models might be ephemeral for very mathematized modelling fields of science such as epidemiology and virology that developed over time very realistic models that enables the possibility of learning with parameters of similar viruses (i.e. SARS) that can definitely help and instruct decision makers to take actions before it is too late.

Here we showed that countries’ importance in the global transportation network has a key role on COVID-19 growth rates in its exponential phase. Our results reinforce board control measures in international airports [42, 43] during exceedingly early stages of pandemics to prevent secondary transmissions that could lead to undesired scenarios of rapid synchronically spread of infectious diseases in different countries. The rapid international spread of the severe acute respiratory syndrome (SARS) from 2002 to 2003 led to extensively assessing entry screening measures at international borders of some countries [44, 45]. The 2019–2020 world spread of COVID-19 highlights that improvements and testing of board control measures (i.e. screening associated with fast testing and quarantine of infected travellers) might be a cheap solution for humanity in comparison to health systems breakdowns and unprecedented global economic crises that the spread of infectious disease can cause. However, it is important to note that board control of potentially infected travellers and how to effectively identify them is still a hotly debated topic in epidemiology and there is still no consensus on accurate methodologies for its application [46].

We do not expect that our results using a macroecological approach at a global scale would have a definitive effect on decision-making in terms of public health in any particular country, province, or city. However, we expect that our analyses show that current claims that growth of COVID-19 pandemics may be lower in developing tropical countries should be taken very carefully, at risk to disturb well-established and effective policy of social isolation that may help to avoid higher mortality rates due to collapse in national health systems.

## Data Availability

All epidemiological, climatic, socioeconomic and transport data used in our study is available to download in online databases.

## Acknowledgments

This paper was developed in the context of the human macroecology project on the National Institute of Science and Technology (INCT) in Ecology, Evolution and Biodiversity Conservation, supported by CNPq (grant 465610/2014–5) and FAPEG (grant 201810267000023). JAFDF, RD, LCT are also supported by CNPq productivity scholarships. We thank Thiago F. Rangel, André Menegotto, Robert D. Morris and Marcus Cianciaruso for their constructive comments on early version of the manuscript.

---

**Fig S1.**
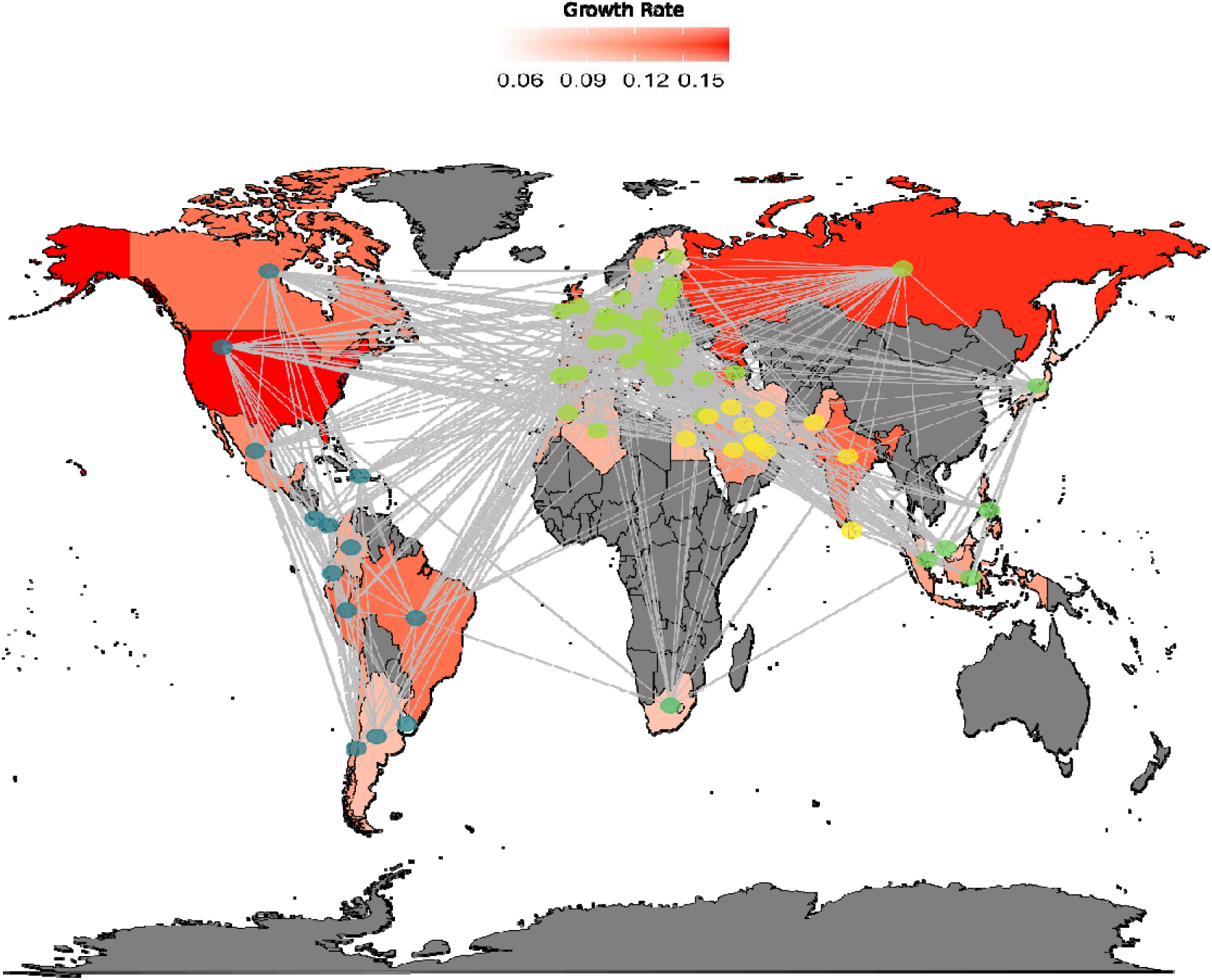
Spatial pattern of the air transportation network among 65 countries that had more than 100 cases and for which time series data had at least 30 days after the 100^th^ case. Red colours in different countries represent different growth rates of COVID-19 in each country. Different colours in the nodes represent modules of countries that are more connected to each other

